# Program Directors’ Perspectives on Duty Hours Following the Physicians’ Work Style Reforms in Japan

**DOI:** 10.1101/2024.02.09.24302425

**Authors:** Kiyoshi Shikino, Yuji Nishizaki, Kazuya Nagasaki, Hiroyuki Kobayashi, Koshi Kataoka, Taro Shimizu, Yasuharu Tokuda

## Abstract

**Background:** In anticipation of Japan’s physician work style reforms scheduled for April 2024, which will restrict physicians’ overtime duty hours to under 960 (Level A) or 1920 (Level C-1) hours per year, this study evaluates the potential impact on clinical resident physicians’ training environments through the lens of residency program directors.

**Methods:** A cross-sectional survey was administered to program directors from 701 facilities associated with the 2023 General Medicine-In Training Examination (GM-ITE), capturing responses regarding their planned level of overtime, perceptions of optimal working hours for training efficacy, and items considered as duty hours.

**Results:** Out of the 701 facilities, 254 responded (36.2% response rate), with a predominant inclination towards adopting Level A (83.5%). A consensus emerged favoring 40 overtime duty hours per month as optimal, significantly lower than previous suggestions of 60–65 hours per week. The study also delineated the classification of activities as duty hours, with patient procedures and mandatory educational sessions identified as such, while self-improvement was largely not considered duty.

**Discussion:** The findings indicate a substantial shift from previously recommended optimal duty hours towards a more conservative approach. This shift may reflect a proactive adaptation to upcoming regulatory changes and raises questions about the potential impact on the quality of clinical training. The distinction between educational activities and duty hours underscores the need for clarity within the impending work style reforms.

## Introduction

Japan’s upcoming physicians’ work style reforms, commencing in April 2024,^1^ establishes limits on overtime duty hours to improve work-life balance. They aim to limit overtime duty hours for all physicians to under 960 hours per year (80 hours per month, Level A), or under 1920 hours per year (160 hours per month, Level C-1).^2^ Previous research indicates that 60–65 duty hours per week (80–100 overtime duty hours per month) may provide the optimal balance for residents’ education, well-being, and patient safety.^3,4,5^ This study assessed the impact of physician’s work style reforms on clinical resident physicians’ environments, in Japan by examining residency program directors’ perceptions of optimal overtime duty hours toward training efficacy and trainee well-being.

## Methods

This cross-sectional study utilized a survey to analyze the responses of program directors of residency training hospitals (701 facilities) that applied for the 2023 General Medicine-In training Examination (GM-ITE). Approximately half of all resident physicians took the GM-ITE in Japan.^5,6^ The survey was conducted from October 18 to December 15, 2023, and included items like the level planned at their own facility, items that fall under working hours rather than self-improvement, and the residency program directors’ perceptions on the optimal working hours for resident physicians. Overtime duty hours per month was divided into eight categories (C1 to C8) (Supplementary file 1).^2,3^

This study was approved by the Ethics Review Board of the Japan Institute for Advancement of Medical Education Program. All participants provided written consent to participate. We adhered to the STROBE reporting guidelines.

## Results

Responses were received from 254 (233 community and 21 university hospitals) of the 701 surveyed facilities (response rate of 36.2%). The implementation of new regulations for resident physicians’ overtime duty hours prompted the majority of the surveyed medical institutions to consider adopting Level A (83.5%). The approach to promoting a balance between the development of clinical skills and the sustenance of physical and mental health revealed a consensus towards 40 hours per month as the optimal amount (22.3% community hospital and 42.8% university hospital) (Figure 1). Of the institutions, 32.7% fell into C2 (20 or more, but less than 40 overtime duty hours per month), and 31.5% fell into C3 (40 or more, but less than 60 overtime duty hours per month) (Supplementary file 2). Most program directors identified patient procedures and mandatory study groups and conferences as duties, rather than self-improvement activities, in contrast to independent study time, which only 2.8% considered as duty (Figure 2).

**Figure 1.**
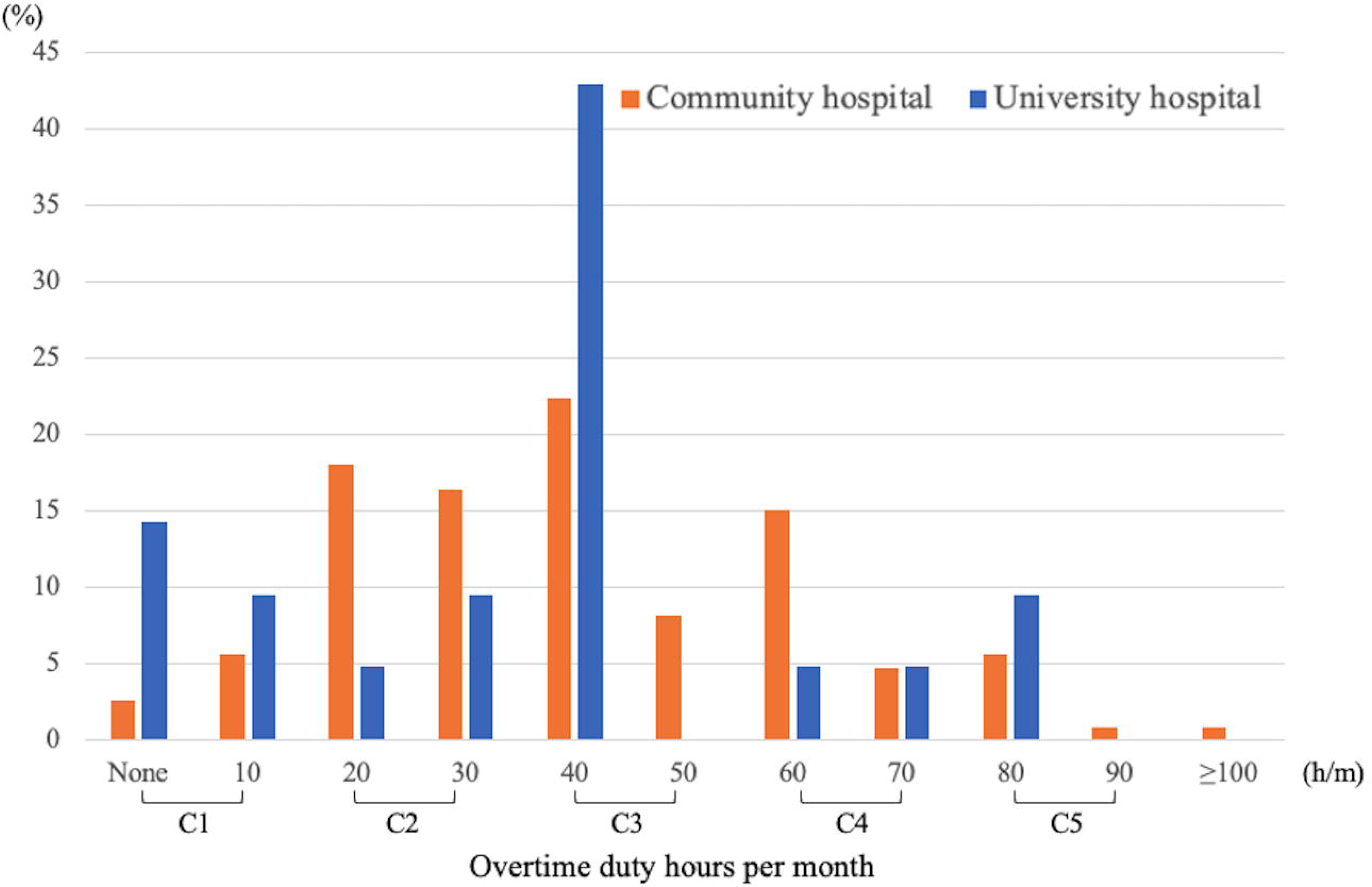
Optimal Monthly Overtime Duty Hours per Month for Resident Physicians by Residency Program Directors. This figure represents the residency program directors’ response on: “*Considering the balance between the development of clinical skills and mental well-being, what do you think is the optimal amount of overtime duty hours per month for resident physicians?”*

**Figure 2.**
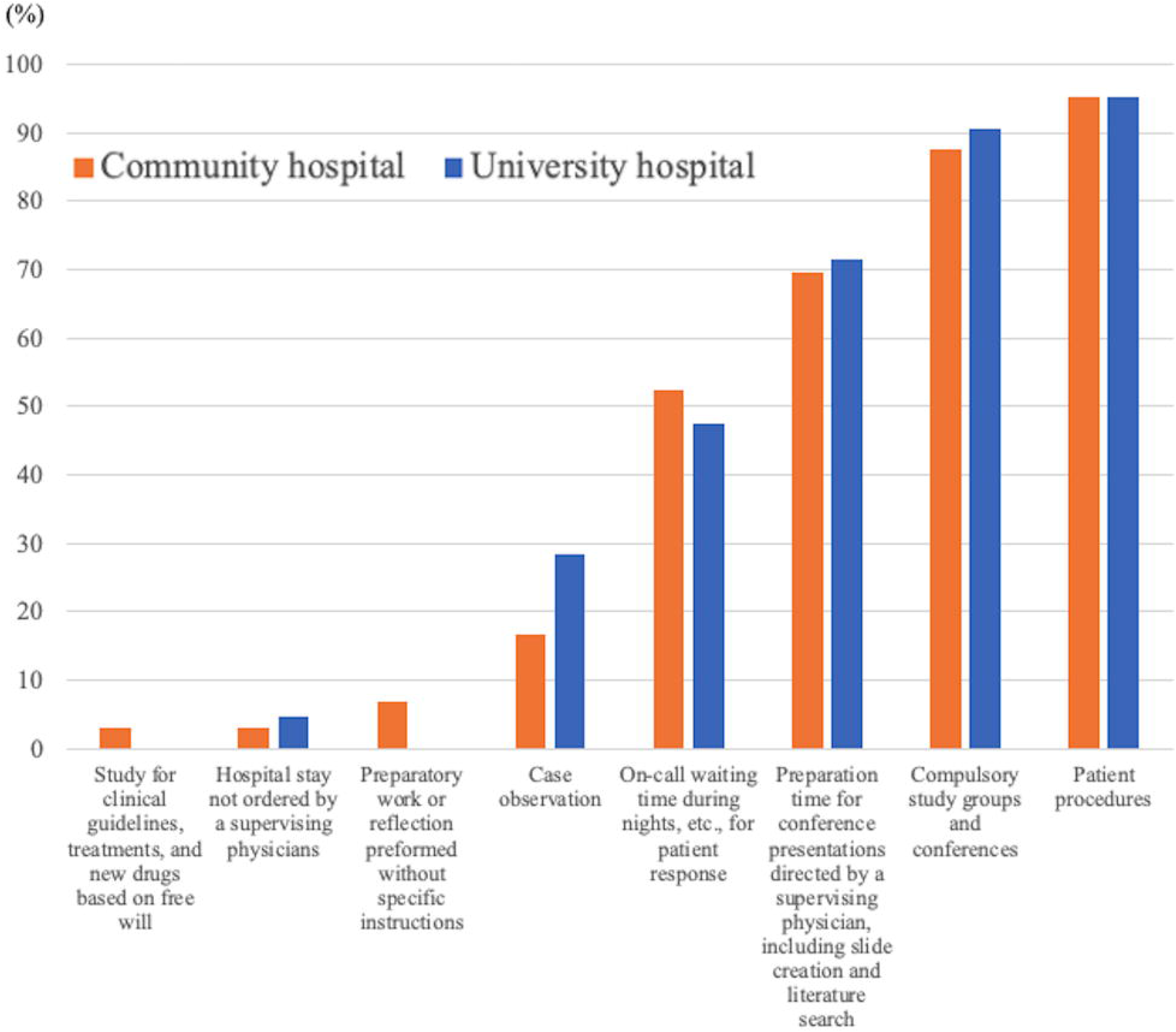
Activities Recognized as “Duty Hours” Rather than “Self-improvement” by Program Directors. This figure represents program directors’ responses on: “*Please select all that apply, which are considered as working hours, not self-improvement*.” (multiple choice)

## Discussion

Despite previous studies suggesting optimal overtime duty hours as 80–100 per month (C5) for resident physicians,^3,4,5^ our survey revealed that residency program directors now lean towards a much lower limit of 40 hours per month (C3), highlighting a significant evidence-practice gap. This conservative shift, likely influenced by the impending reforms and anticipation of stricter overtime enforcement, suggests a preemptive adjustment for compliance, and also raises concerns regarding the quality of clinical training. Moreover, the clear recognition of patient procedures and compulsory educational activities as duty hours, as opposed to self-improvement, underscores the complexity of delineating educational activities within their limits.

The study’s response rate was 36.4%, and did not encompass all 1037 clinical training hospitals in Japan, which may limit the generalizability of the results.

While many residency training hospitals implemented Level A limits, residency program directors considered that shorter limits were appropriate for their resident physicians. A concerted effort to bridge the gap between policy-, practice-, and evidence-based standards to maximize the development of clinical expertise of the resident physicians is crucial for successfully enacting these reforms.

## Supporting information

Supplementary file 2. Optimal Monthly Overtime Duty Hours per Month for Resident Physicians by Residency

Supplementary file 3. Data Sharing Statement

## Data Availability

Data from the GM-ITE can be made available to researchers with ethical permissions to access that data for the specified purposes.

## Figure Legends

**Supplementary file 1. Category of Physicians’ Duty Hours**

Duty hours were determined by surveying the average total weekly duty hours. Average duty hours were calculated by summing the hours spent on weekdays, on-call duties, and weekend duties. On-call standby time (constrained time) was also included in duty hours. For these calculations, it was assumed that one month consisted of four weeks.

**Supplementary file 2. Optimal Monthly Overtime Duty Hours per Month for Resident Physicians by Residency**

This table represents the residency program directors’ response on: “*Considering the balance between the development of clinical skills and mental well-being, what do you think is the optimal amount of overtime duty hours per month for resident physicians?”*

**Supplementary file 3. Data Sharing Statement**

