## Supplemental Data 1 for "Program Directors’ Perspectives on Duty Hours Following the Physicians’ Work Style Reforms in Japan"

**Supplementary file 1. Category of Physicians’ Duty Hours**

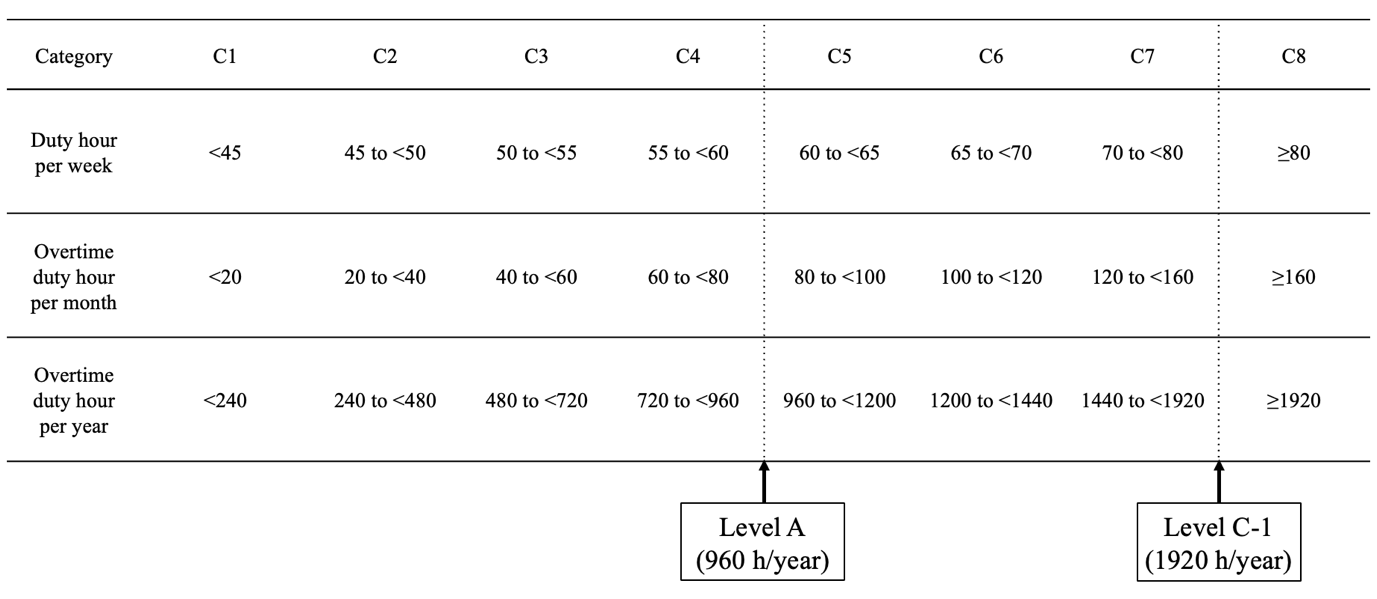

Duty hours were determined by surveying the average total weekly duty hours. Average duty hours were calculated by summing the hours spent on weekdays, on-call duties, and weekend duties. On-call standby time (constrained time) was also included in duty hours. For these calculations, it was assumed that one month consisted of four weeks.
