## Supplementary file 2. Optimal Monthly Overtime Duty Hours per Month for Resident Physicians by Residency for "Program Directors’ Perspectives on Duty Hours Following the Physicians’ Work Style Reforms in Japan"

| Overtime duty hour per month | None | 10 | 20 | 30 | 40 | 50 | 60 | 70 | 80 | 90 | ≥100 |
| --- | --- | --- | --- | --- | --- | --- | --- | --- | --- | --- | --- |
| Total (n, %) | 9  (3.5) | 15 (5.9) | 43 (16.9) | 40 (15.8) | 61 (24.0) | 19 (7.5) | 36 (14.2) | 12 (4.7) | 15 (5.9) | 2 (0.8) | 2 (0.8) |
| Community  hospital (n, %) | 6 (2.6) | 13 (5.6) | 42 (18.0) | 38 (16.3) | 52 (22.3) | 19 (8.1) | 35 (15.0) | 11 (4.7) | 13 (5.6) | 2 (0.9) | 2 (0.9) |
| University hospital (n, %) | 3 (14.3) | 2 (9.5) | 1 (4.8) | 2 (9.5) | 9 (42.8) | 0 (0) | 1 (4.8) | 1 (4.8) | 2 (9.5) | 0 (0) | 0 (0) |

This table represents the residency program directors’ response on: “*Considering the balance between the development of clinical skills and mental well-being, what do you think is the optimal amount of overtime duty hours per month for resident physicians?”*
