## Supplementary file 3. Data Sharing Statement for "Program Directors’ Perspectives on Duty Hours Following the Physicians’ Work Style Reforms in Japan"

**Data**

**Data available:** No

**Additional Information**

**Explanation for why data not available:** Data from the GM-ITE can be made available to researchers with ethical permissions to access that data for the specified purposes.
